# Improving Individualized Treatment Decisions: A Bayesian Multivariate Hierarchical Model for Developing a Treatment Benefit Index using Mixed Types of Outcomes

**DOI:** 10.1101/2023.11.17.23298711

**Authors:** Danni Wu, Keith S. Goldfeld, Eva Petkova, Hyung G. Park

## Abstract

**Background:** Precision medicine has led to the development of targeted treatment strategies tailored to individual patients based on their characteristics and disease manifestations. Although precision medicine often focuses on a single health outcome for individualized treatment decision rules (ITRs), relying only on a single outcome rather than all available outcomes information leads to suboptimal data usage when developing optimal ITRs.

**Methods:** To address this limitation, we propose a Bayesian multivariate hierarchical model that leverages the wealth of correlated health outcomes collected in clinical trials. The approach jointly models mixed types of correlated outcomes, facilitating the “borrowing of information” across the multivariate outcomes, and results in a more accurate estimation of heterogeneous treatment effects compared to using single regression models for each outcome. We develop a treatment benefit index, which quantifies the relative treatment benefit of the experimental treatment over the control treatment, based on the proposed multivariate outcome model.

**Results:** We demonstrate the strengths of the proposed approach through extensive simulations and an application to an international Coronavirus Disease 2019 (COVID-19) treatment trial. Simulation results indicate that the proposed method reduces the occurrence of erroneous treatment decisions compared to a single regression model for a single health outcome. Additionally, the sensitivity analysis demonstrates the robustness of the model across various study scenarios. Application of the method to the COVID-19 trial exhibits improvements in estimating the individual-level treatment efficacy (indicated by narrower credible intervals for odds ratios) and optimal ITRs.

**Conclusion:** The study jointly models mixed types of outcomes in the context of developing ITRs. By considering multiple health outcomes, the proposed approach can advance the development of more effective and reliable personalized treatment

## 1 Introduction

In recent years, the growing emphasis on tailoring treatment strategies for patients according to their unique characteristics and disease manifestations has fueled a surge of interest among researchers and clinicians in the development of individualized treatment decision rules (ITRs) [1–12]. A significant challenge in developing robust and accurate ITRs is in handling noisy outcome data. Typical methods for developing ITRs rely solely on a single health outcome, thus limiting the full exploitation of the available outcomes data. This limitation can lead to suboptimal data usage for individualized treatment decision-making, subsequently yielding a considerable degree of uncertainty, particularly when the outcome data are noisy.

To address this issue, we capitalize on the wealth of correlated and clustered health outcomes collected in trials by utilizing multivariate models, which have demonstrated significant improvements in estimation and prediction accuracy compared to their univariate counterparts [13–19]. Although correlated and clustered observations are often modeled (in the frequentist paradigm) by a marginal model via generalized estimating equations or a generalized linear mixed model [20], Bayesian methods can handle highly complex hierarchical structures and efficiently estimate parameters via Markov Chain Monte Carlo sampling, making it an appealing and efficient strategy [21–23].

We propose a Bayesian multivariate hierarchical model that explicitly accounts for patient heterogeneity and enables the “borrowing of information” among multiple correlated mixed types of outcomes, resulting in a more accurate estimation of treatment effects. Based on the proposed model, we employ a treatment benefit index [24, 25] to optimize ITRs.

Existing methods for ITRs in the presence of multiple outcomes have been proposed [26–36], including estimation of composite outcomes [34, 35], estimating patients’ outcome preferences [31, 33, 37], “set-valued” approaches [27, 28] and constrained estimation [26, 30] that focuses on balancing competing outcomes. Our approach distinguishes itself by focusing on improving the modeling efficiency and building the connection between correlated mixed types of outcomes through a Bayesian hierarchical model, which allows the treatment effects for each outcome to share a common prior distribution. This strategy is particularly effective when there is reason to believe that the treatment exerts similar influences on the outcomes. By effectively accommodating dependency in multiple highly correlated health outcomes, our approach improves the estimation of treatment effects at both the patient and outcome-specific levels. Simulation results demonstrate the substantial gains in performance offered by the proposed Bayesian multivariate hierarchical model. Our method is applied to a clinical trial of Coronavirus disease 2019 (COVID-19) convalescent plasma treatment. In the Continuous Monitoring of Pooled International Trials of Convalescent Plasma for COVID-19 Hospitalized Patients (COMPILE) trial [38, 39], multiple correlated health outcomes were collected, including the primary ordinal outcome measure [40] and several secondary outcomes. However, the model’s applicability extends beyond this specific case, serving as a versatile tool for analyzing mixed types of outcome data and developing ITRs in clinical trials. By providing enhanced estimations of heterogeneous treatment effects and more accurately quantified uncertainty measurements reflecting all the available information from multiple health outcomes, this innovation holds the potential to guide clinical practice and contribute to the optimization of individualized treatment strategies.

We organize the paper as follows. In the Methods section, we present the Bayesian multivariate model for evaluating heterogeneous treatment effects and developing ITRs. We elucidate the reasoning behind our selection of prior distributions. In the Results section, we present extensive simulation results, which enabled us to compare the performance of the proposed multivariate model for mixed types of outcomes with a Bayesian single regression model for a single outcome. Our methodology yields more precise estimations of heterogeneous treatment effects and reduces the occurrence of erroneous optimal treatment decisions. Then, we validate the robustness of our proposed model through sensitivity analysis. We have applied this model to data from an international COVID-19 study, COMPILE, demonstrating its ability to provide more accurate estimations of heterogeneous treatment effects, as represented by odds ratios (ORs) with narrower credible intervals (CrIs) reflecting all available outcomes information. In the Discussion and conclusion section, we provide a discussion and offer insights into potential future applications of our work.

## 2 Methods

In this section, we present a Bayesian approach for modeling mixed types of outcomes within the exponential family. Let ***Y*** _*i*_ represent the vector of outcomes of length *d* for the *i*^*th*^ subject (*i* = 1,…, *n*), where the *k*^*th*^ element 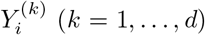 follows an exponential family distribution. We denote ***η*** = (***η***_1_,…, ***η***_*n*_), with 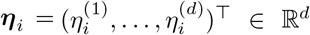, and 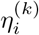 as the canonical parameter associated with the assumed distribution of 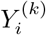. Additionally, we define ***ϕ*** = (*ϕ*^(1)^,…, *ϕ*^(*d*)^)^⊤^ ∈ ℝ^*d*^, where *ϕ*^(*k*)^ *>* 0 is an unknown dispersion parameter.

Conditional on ***η*** _*i*_ and ***ϕ***, the *d* components of 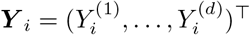 are assumed to be independent. The likelihood of ***Y*** = (***y***_1_,…, ***y***_*n*_) can be expressed as:

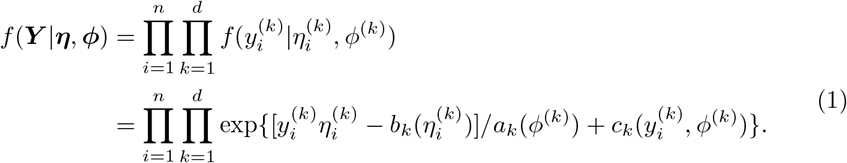

Here, the functions *a*_*k*_(·), *b*_*k*_(·), and *c*_*k*_(·) are exponential family distribution-specific known functions for the *k*^*th*^ outcome, whereas 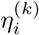 and *ϕ*^(*k*)^ are unknown quantities.

In the model, we relate the outcome-specific expected value with the linear combination of covariates and treatment assignment via a canonical parameter *η*^(*k*)^ and an outcome-specific canonical link *g*^(*k*)^(.) depending on the type of outcomes (e.g., identity function for continuous outcomes, logit function for binary outcomes, and log function for count outcomes):

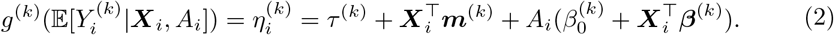

Here, *τ* ^(*k*)^ is the outcome-specific intercept, ***m***^(*k*)^ is the length-*p* main effect of the pre-treatment characteristics ***X***_*i*_ on the *k*^*th*^ outcome, 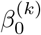 is the main effect of the experimental treatment (*A* = 1) on the *k*^*th*^ outcome, and ***β***^(*k*)^ is the length-*p* ***X***-by-*A* interaction effect on the *k*^*th*^ outcome.

For patients with pre-treatment characteristics ***x***, the treatment-control effect contrast based on (2) is defined as:

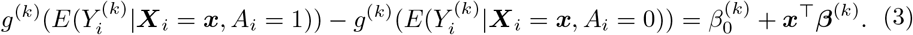

This treatment-control effect contrast is the primary focus in clinical trials. For example, if the outcome is binary and *g*^(*k*)^(.) is a *logit* link function, 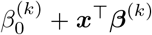 corresponds to the effect of the experimental treatment vs. control on the *k*^*th*^ outcome, as measured by the log odds ratio (log OR). Assuming that the first outcome (*k* = 1) is the primary outcome and a lower value of this outcome is preferable. Then, a log OR below 0 signifies that the experimental treatment yields a more favorable primary outcome compared to the control treatment. Equation (3) demonstrates that the treatment-control effect contrast, e.g. log OR, depends solely on the main effect of treatment *A* and the ***X***-by-*A* interaction effects and not on the ***X*** main effects. Our Bayesian model’s objective is to efficiently estimate the effect of treatment *A* (represented by 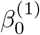 in Equation (3)) and the ***X***-by-*A* interaction effects (represented by ***β***^(1)^ in Equation (3)) on the primary outcome *Y* ^(1)^ by “borrowing information” from other correlated outcomes *Y* ^(*k′*)^, *k*^′^ *>* 1.

### 2.1 Individualized treatment decision rule

Our goal is to predict optimal treatment options for future patients, taking into account their pre-treatment characteristics. Bayesian analysis can provide the full posterior distribution of the parameter of interest. We define the treatment benefit index (TBI) for a patient with pre-treatment characteristics ***x*** as the posterior probability that the treatment-control contrast in Equation (3) is less than 0:

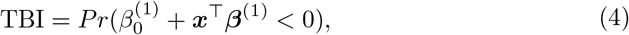

representing the (posterior) probability that the experimental treatment is more beneficial than the control treatment. The estimated optimal ITR, denoted as *â*_*opt*_: *x* ↦ {0, 1}, is defined based on the TBI in (4):

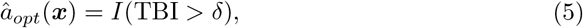

where *I*(.) is the indicator function, and 0 *< δ <* 1 is a threshold probability to make treatment decisions. We set the threshold *δ* to 0.5 in this paper. If the TBI exceeds 0.5, then the patient is recommended to receive the experimental treatment (i.e., *â*_*opt*_(***x***) = 1), as there is a more than 0.5 probability that the experimental treatment is more beneficial than the control treatment.

### 2.2 Model and prior specification

In this section, we describe a versatile framework for modeling mixed types of outcomes. The framework was motivated by the COMPILE study, in which we encountered the need to jointly model a primary ordinal outcome and binary outcomes. Although we demonstrate the applicability and utility of our proposed framework using ordinal and binary outcomes as an example, the framework is designed to be adaptable to other mixed outcome types.

To model the primary ordinal outcome, a cumulative proportional odds (*co*) model was determined to be the most appropriate method [41]. Let *Y* ^(1)^ represent the *L* levels ordinal outcome, with probabilities 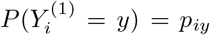 for *y* = 1,…, *L*. The cumulative probabilities are modeled as 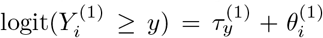, where 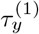’s represent the intercepts for *y* = 2,…, *L* and satisfy the monotonicity requirement for the intercepts of the *co* model, and 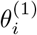 is a linear predictor defined below. Logistic models are used to analyze the binary outcomes. Let *Y* ^(2)^,…, *Y* ^(*d*)^ denote the *d* − 1 binary outcomes. Bernoulli distributions with probabilities 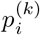 are assumed, such that 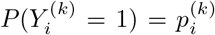 for *k* = 2,…, *d*. The probabilities are modeled as 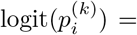 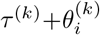, where *τ* ^(*k*)^ are the intercepts and 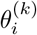 are the linear predictors defined below. For each unit of analysis *i* ∈ {1,…, *n*}, a binary treatment assignment *A*_*i*_ ∈ {0, 1} is considered, with *A*_*i*_ = 1 representing the experimental treatment and *A*_*i*_ = 0 denoting the control treatment. A multivariate outcome 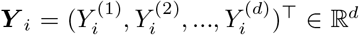 and a vector of pre-treatment characteristics ***X***_*i*_ ∈ ℝ^*p*^ are taken into account.

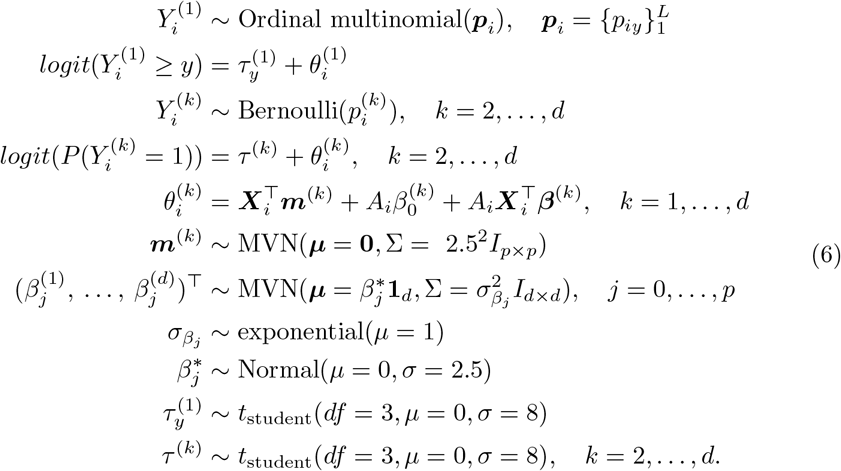

It is crucial to identify prior distribution assumptions in Bayesian statistics. We followed the criteria described in [42] for selecting prior distributions.

#### Outcome-specific treatment main effect 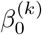 and interaction effect *β*^(*k*)^

To facilitate flexible information sharing about the coefficients across outcomes, we employ hierarchical shrinkage. The prior distribution assumes that each outcome-specific treatment main effect 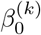 is closely centered around a pooled “treatment main effect” 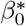, postulating that these treatment effects are comparable across all outcomes. The variation of each outcome-specific treatment main effect around the group’s mean 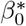 is represented by its standard deviation 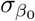. The outcome-specific interaction effect 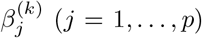 is distributed as 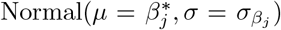, where 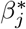 denotes the pooled “interaction effect” across all outcomes. 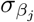 controls the strength of information borrowing. A large mean of the prior distribution of 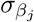 allows for greater variability, whereas a small value constrains the coefficients to remain closer to the pooled effect. In the Simulation illustration section, we assigned a prior mean of 1 to 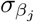.

For the outcome-specific intercepts *τ* ^(*k*)^, we use a *t*_student_ distribution with 3 degrees of freedom (*σ* = 8). This choice offers heavier tails compared to the *Normal* distribution (*σ* = 8), ensuring that the Hamiltonian Monte Carlo (HMC) sampling [43] has adequate flexibility for exploring the sample space. In the case of covariates’ main effects ***m***^(*k*)^, we use a diffuse prior, with the expectation that the observed data will primarily drive the shape of the posterior distribution. Similarly, for the pooled treatment main effect and interaction effects across outcomes, 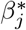, we adopt a diffuse prior.

## 3 Results

In this section, we present a comparative analysis of two Bayesian models for estimating heterogeneous treatment effects and ITRs. Specifically, the performance of the proposed multivariate model will be compared to that of a univariate model, which only relies on a single primary outcome. We also conducted sensitivity analyses to assess the robustness of the proposed model across different study scenarios. We then applied our method to an international COVID-19 clinical trial and examined the proposed model for goodness-of-fit.

### 3.1 Simulation illustration

We conducted a series of simulation experiments. The Bayesian models were implemented using Stan [43], which enables Bayesian inference based on HMC, with the No-U-Turn sampler[43].

#### 3.1.1 Simulation setup and performance evaluation

We used the R package *simstudy* [44] to generate simulated data sets. For a given sample size in the training dataset, denoted as *n*, we independently generated treatment indicators, denoted *A*_*i*_ ∈ {0, 1}, from the Bernoulli distribution with a probability of *P* (*A*_*i*_ = 1) = 0.5. The covariates ***X***_*i*_ ∈ ℝ^*p*^ comprised 3 independent binary variables generated from the Bernoulli distribution with probability *P* (*X*_*i*_ = 1) = 0.5, and *p* − 3 independent continuous variables, drawn from the multivariate normal distribution with mean zero and unit variance. We generated a set of four outcomes 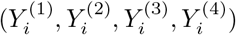, mimicking the outcomes collected from the COMPILE study. The variable 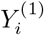 follows an 11-level ordinal multinomial distribution, while 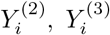, and 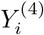, representing the 3 supplementary binary outcomes, are generated using Bernoulli distributions. The true parameter values used for the data generation are as follows. These notations adhere to model (6). We consider *p* = 5 covariates, and the covariates’ main effect coefficients for each of the 4 outcomes are ***m***^(1)^ = [0.35, −0.40, 0.15, 0.20, −0.21]^⊤^, ***m***^(2)^ = [0.40, −0.38, 0.13, 0.19, −0.22]^⊤^, ***m***^(3)^ = [0.38, −0.39, 0.14, 0.18, −0.20]^⊤^, ***m***^(4)^ = [0.42, −0.41, 0.16, 0.21, −0.19]^⊤^.

• Treatment’s main effect coefficient for each outcome:
  - 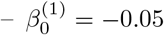
  - 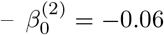
  - 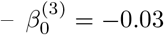
  - 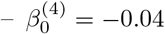
***X***-by-*A* interaction effect coefficients for each outcome:
  - ***β***^(1)^ = [0.20 −0.10 0.10 0.05 −0.06]^⊤^
  - ***β***^(2)^ = [0.19 −0.11 0.09 0.04 0.07]^⊤^
  - ***β***^(3)^ = [0.18 −0.12 0.11 0.06 −0.05]^⊤^
  - ***β***^(4)^ = [0.21 −0.09 0.12 0.07 −0.04]^⊤^

As a comparison model for model (6), we employed a Bayesian univariate model (7) that only uses the single primary ordinal outcome, specified as follows:

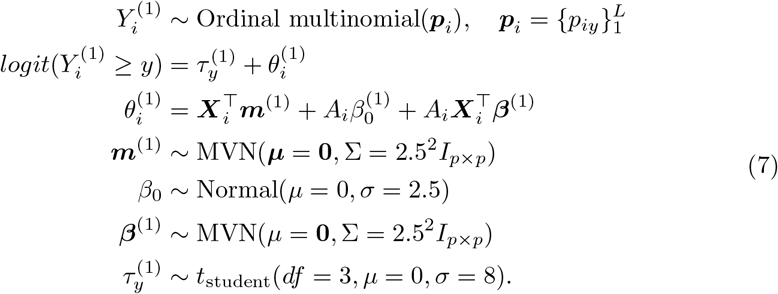

As evaluation metrics for the performance of the models, we consider two criteria: the proportion of correct decisions (PCD) and the Area Under the Receiver Operating Characteristic (ROC) curve. The PCD corresponds to the proportion of cases (*i* = 1,…, *ñ*, and *ñ* = 2000 is the testing set sample size) with *â*_*opt*_(***x***_*i*_) = *a*_*opt*_(***x***_*i*_), where *â*_*opt*_(***x***_*i*_) is defined in Equation (5) with threshold *δ* = 0.5, and the true optimal ITR *a*_*opt*_(***x***_*i*_) = *I*(OR *<* 1), where 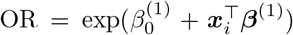, with 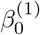 and 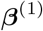 corresponding to the true values used in the data generation process. Since without loss of generality, we assume a lower value of the outcome indicates a better health condition, an OR less than 1 indicates the experimental treatment is better than the control treatment.

PCD is computed using a decision threshold *δ* = 0.5 as per Equation (5). Another evaluation metric is the Area Under the ROC Curve (AUC), which does not rely on the selection of a specific decision threshold, and accounts for the trade-off between true positive rate (sensitivity) and false positive rate (1 - specificity) for various decision thresholds. AUC values range from 0 to 1, with a higher value indicating a better classification performance [45]. To calculate the AUC, we first evaluate the estimated TBI as defined in Equation (4) on the test data, and then generate the ROC curve, considering every unique TBI value as a potential threshold; for each threshold, we compute *â*_*opt*_(***x***) according to Equation (5), and compare it with *a*_*opt*_(***x***) to calculate the true positive and false positive rates. Then the auc function from the *pROC* package [46] is used to compute the AUC.

We present simulation results for various training sample sizes, *n* ∈ {250, 500, 1000, 2000}, and a fixed test dataset size of 2000. For each *n*, we conducted 1000 simulations, with each simulation using 2000 HMC iterations for warm-up and retaining 10000 iterations for inference.The Stan code for the Bayesian multivariate hierarchical model is provided in Additional file 1.

The plot in Figure 1 presents a comparison of the performance of the multivariate model (6) and the univariate model (7) based on their PCD and AUC values. The performance is evaluated across varying training set sizes, represented by the number of subjects in the training set on the x-axis. The y-axis displays the PCD or AUC values, with higher values indicating better model performance. The figure illustrates that the multivariate model (in orange) generally exhibits higher PCD and AUC values compared to the univariate model (in blue) across all training set sizes, suggesting that the proposed multivariate model outperforms its univariate counterpart with respect to making correct treatment decisions for subjects in the testing set.

**Fig. 1.**
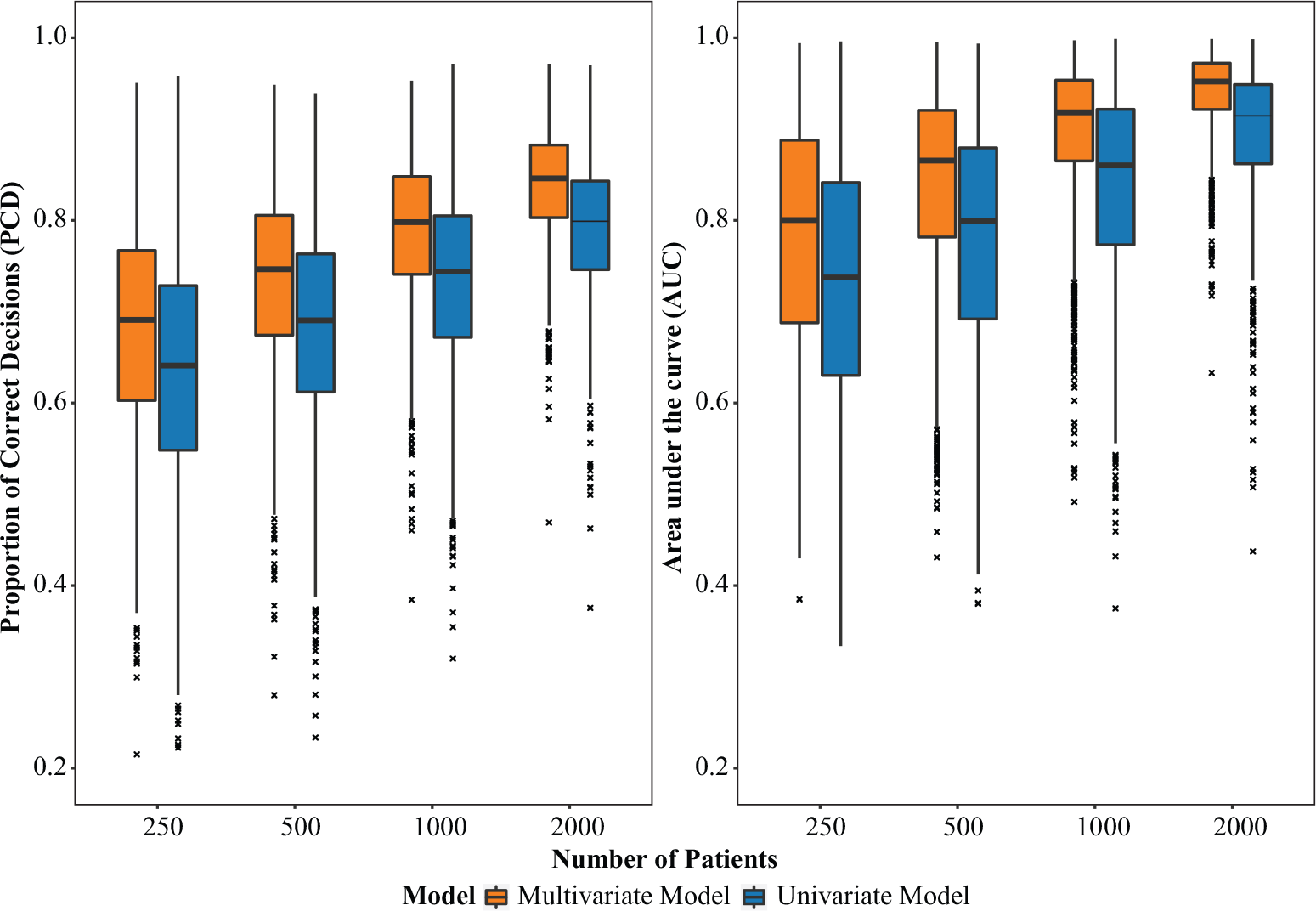
Boxplots of the proportion of correct decisions (PCD) and area under the curve (AUC) in the test sets, comparing the multivariate (orange) and univariate (blue) models across different training set sizes (as indicated in the x-axis). Each box shows the interquartile range (IQR), with the horizontal line inside the box representing the median PCD and AUC value. The whiskers extend to the minimum and maximum PCD and AUC values within 1.5 times the IQR. Outliers are represented by small cross symbols.

Some experts believe that the true optimal ITR should be based on potential outcomes. In light of this perspective, we also provide a comparison of the performance of the Bayesian multivariate and univariate models utilizing the new potential outcomes-based ITR in Additional file 2. Despite the less remarkable improvement in PCD and AUC, our proposed model (6) still outperforms the univariate model (7).

#### 3.1.2 Sensitivity analysis

Each patient’s individual-level treatment efficacy for a specific outcome can vary, making it logical to incorporate random effects into the data generation process. In this section, we conducted a sensitivity analysis to assess the robustness of the models. To simulate various study scenarios, we introduce a modified data generation model that incorporates additional parameters, *γ*_*i*0_ and **Γ**_*i*_:

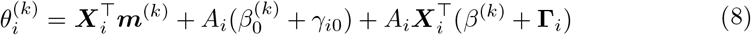

The *γ*_*i*0_ indicates the random effect associated with treatment, and **Γ**_*i*_ indicates the random effect associated with ***X***-by-*A* interaction. The standard deviation for both parameters is determined by *σ*. The true values of the other parameters follow the data generation process described in Section Simulation setup and performance evaluation. We considered a range of values for *σ* ∈ {0.1, 0.2, 0.3}, as well as different training sample sizes *n* ∈ {250, 500, 1000, 2000}, with a fixed test dataset size of 2000. For each set of *σ* and *n*, we conducted 1000 simulations. The PCD and AUC are presented in Figure 2. In the plot, the y-axis represents PCD or AUC, while the x-axis displays the number of subjects in the training set. The multivariate model (6) consistently outperforms the univariate model (7). However, when *σ* = 0.2 and 0.3, the superiority of the multivariate model becomes less pronounced. This is because the true values of the main effect of treatment and fixed effect of the interaction term are all ≤ 0.21, and *σ* = 0.2 and 0.3 already constitute relatively large values of random individual effects. Even with such a relatively large *σ* value, the proposed model (6) still outperforms the univariate model (7), demonstrating the robustness of our approach. Using the same setting of sensitivity analysis, we also provide a comparison of the performance of multivariate model (6) and univariate model (7) utilizing the potential outcomes-based ITR in Additional file 3.

**Fig. 2.**
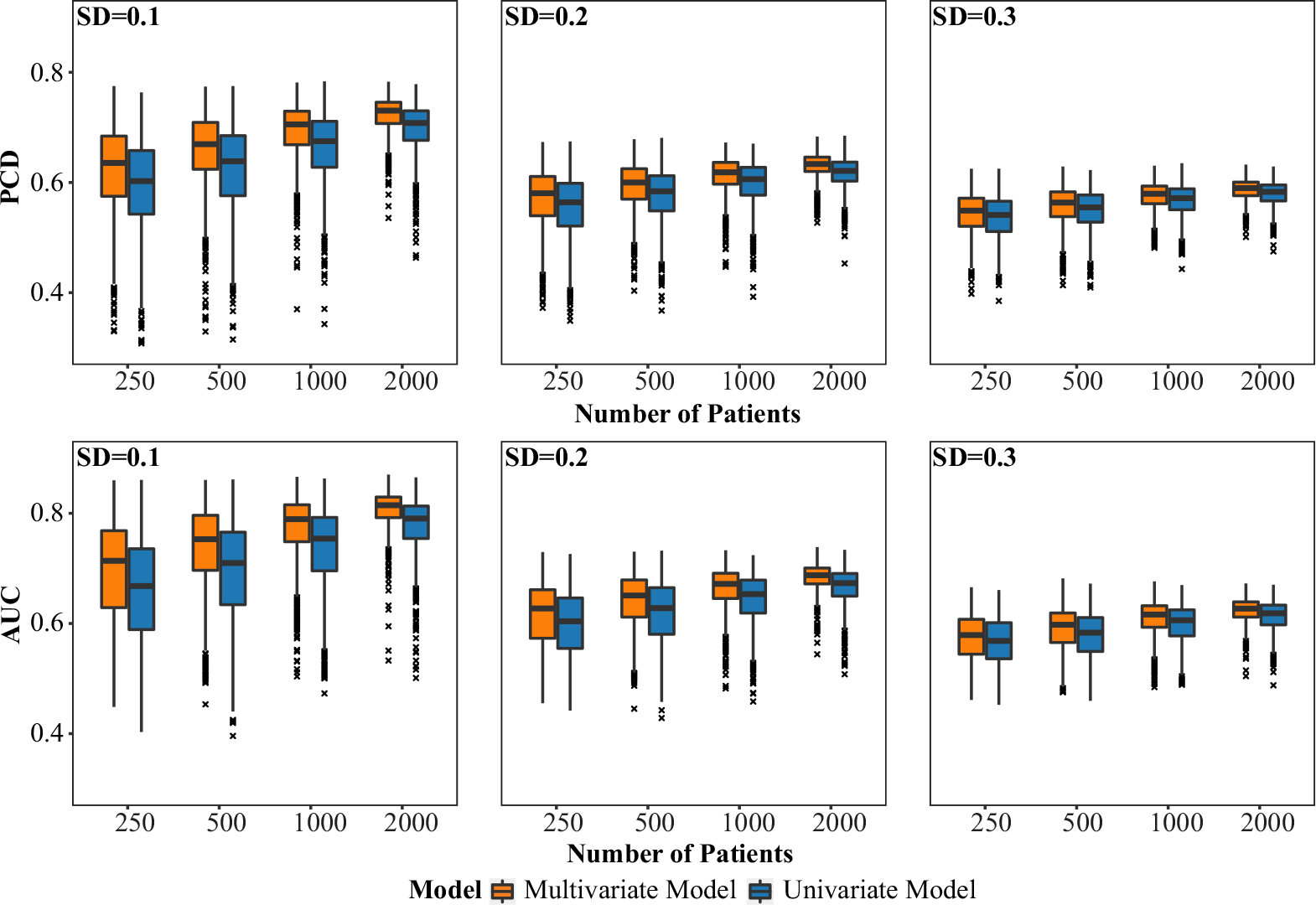
Boxplots of PCD (upper panel) and AUC (lower panel) in the test sets, comparing the multivariate (orange) and univariate (blue) models across different training set sizes (as indicated in the x-axis) and different standard deviations (FSDs) of random effects. Three different levels of SD for random effects are considered in the data generation process: SD=0.1, SD=0.2, and SD=0.3.

### 3.2 Application to data from a COVID-19 randomized clinical trial

In this section, we apply the proposed Bayesian multivariate model to data from 2341 patients in the COMPILE COVID-19 clinical trial, focusing on the CCP treatment for hospitalized COVID-19 patients not on mechanical ventilation at the time of randomization [38, 39, 42]. This study collected several mixed types of outcomes, including a primary outcome and supplementary/secondary outcomes. Park et al. [24] developed an ITR solely based on the primary outcome using a frequentist method. The current paper also focuses on the primary outcome. However, the proposed approach reduces the uncertainty associated with the estimation of heterogeneous treatment effects and ITRs by jointly modeling the mixed types of outcomes using Bayesian techniques and “borrowing information” across correlated outcomes.

The primary outcome is the World Health Organization (WHO) 11-point clinical scale, measured at 14 ± 1 day after randomization (hereafter, day 14), assessing COVID-19 severity with values ranging from 0 (no infection) to 10 (death) [47]. To “borrow information” we employ binary outcomes collected in the COMPILE study, such as hospitalization, ventilation or worse, and death at 28 ± 2 days after randomization (hereafter, day 28). We used the same set of pre-treatment characteristics as in the ITR from Park et al. [24], which was selected via extensive cross-validation. The pre-treatment characteristics are listed below.

- Pre-treatment characteristics in the treatment-by-***X*** interaction effects term: WHO score at baseline (an ordinal variable represents hospitalized but no oxygen therapy required, hospitalized with oxygen required via mask or nasal prongs, and hospitalized with high-flow oxygen required); WHO score at baseline & Age ≥ 67 interaction; Indicator for blood type A or AB; Indicator for the presence of Cardiovascular Disease; Indicator for comorbid Diabetes Mellitus & Pulmonary Disease.
- Pre-treatment characteristics in the main effects term: Age (mean (SD) of 60.3 (15.2) years); Sex (35.7% were women); WHO score at baseline; WHO score at baseline & Age interaction; Indicator for blood type A or AB; Indicator for comorbid Diabetes Mellitus & Cardiovascular Disease interaction; Indicator for comorbid Diabetes Mellitus & Pulmonary Disease interaction; Duration of symptoms before randomization (a binary variable defined as 0-6 days and ≥ 7 days); Quarter during which patient was enrolled (a categorical variable that represents Jan-March 2020, Apr-June 2020, July-Sept 2020, Oct-Dec 2020, and Jan-March 2021); Indicator of treatment (a binary variable with 1 for CCP treatment, and 0 for control treatment).

Our analysis used complete cases, yielding a final sample of 2287 patients (the number of patients at different clinical stages of COVID-19 measured on the WHO 11-point scale at day 14 by treatment group is provided in Additional file 4). We evaluated the performance of the two models: the multivariate outcome model (6) and the univariate outcome model (7). As it is expected that the main effect of treatment 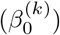 should not vary significantly across different outcomes and interaction effects 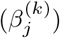 exhibit relatively small variation across outcomes, we employed an informative prior 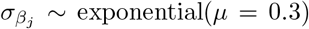 on the hierarchical standard deviation parameter 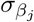. In Figure 3, we presented the posterior distributions (medians and 95% CrIs) of coefficients for treatment and pre-treatment patient characteristics (in terms of *log*OR) associated with the TBI for the primary ordinal outcome from both models, (6) and (7). Table 1 presents the posterior distributions of coefficients for treatment and pre-treatment characteristics (in terms of *log*OR) for all ordinal and binary outcomes.

**Table 1.** The estimated TBI index coefficients (Median [95%CrI]) for treatment and pre-treatment characteristics (*log*OR) under univariate and multivariate models.

| Treatment and pre-treatment characteristics | Index coefficients (Median [95%CrI]) |  |  |  |  |
| --- | --- | --- | --- | --- | --- |
|  | Univariate model<br>Primary ordinal outcome | Multivariate model<br>Primary ordinal outcome | Binary outcome 1 | Binary outcome 2 | Binary outcome 3 |
| Diabetes & Pulmonary | -0.37 [-1.11, 0.37] | -0.51 [-1.05, 0.06] | -0.51 [-1.10, 0.18] | -0.61 [-1.37, 0.00] | -0.65 [-1.56, -0.06] |
| Cardiovascular Disease | -0.26 [-0.51, -0.01] | -0.32 [-0.51, -0.11] | -0.35 [-0.59, -0.11] | -0.37 [-0.67, -0.15] | -0.37 [-0.68, -0.14] |
| Blood Type A or AB | -0.28 [-0.58, 0.03] | -0.37 [-0.62, -0.09] | -0.47 [-0.82, -0.19] | -0.49 [-0.89, -0.20] | -0.43 [-0.78, -0.12] |
| Oxygen by high flow* & Age<sub>≥67</sub> | 0.31 [-0.28, 0.91] | 0.41 [-0.01, 0.81] | 0.46 [0.03, 0.93] | 0.46 [0.03, 0.92] | 0.43 [-0.05, 0.87] |
| Oxygen by mask or nasal prongs* & Age<sub>≥67</sub> | 0.12 [-0.22, 0.45] | 0.05 [-0.22, 0.31] | 0.06 [-0.24, 0.40] | -0.02 [-0.42, 0.28] | -0.01 [-0.42, 0.29] |
| Oxygen by high flow* | 0.24 [-0.31, 0.80] | 0.54 [0.12, 0.94] | 0.56 [0.12, 1.00] | 0.59 [0.15, 1.04] | 0.56 [0.11, 1.01] |
| Oxygen by mask or nasal prongs* | 0.32 [-0.09, 0.72] | 0.67 [0.34, 0.99] | 0.75 [0.40, 1.16] | 0.75 [0.37, 1.16] | 0.71 [0.32, 1.11] |
| CCP Treatment | -0.18 [-0.55, 0.19] | -0.39 [-0.68, -0.11] | -0.41 [-0.73, -0.10] | -0.41 [-0.74, -0.10] | -0.44 [-0.8, -0.13] |

Note: \* The reference level: hospitalized but no oxygen therapy required.
The primary ordinal outcome is the WHO 11-point clinical on day 14.
The supplementary binary outcomes are as follows: (1) hospitalization at day 28, (2) the need for ventilation or worse at day 28, (3) mortality at day 28.

**Fig. 3.**
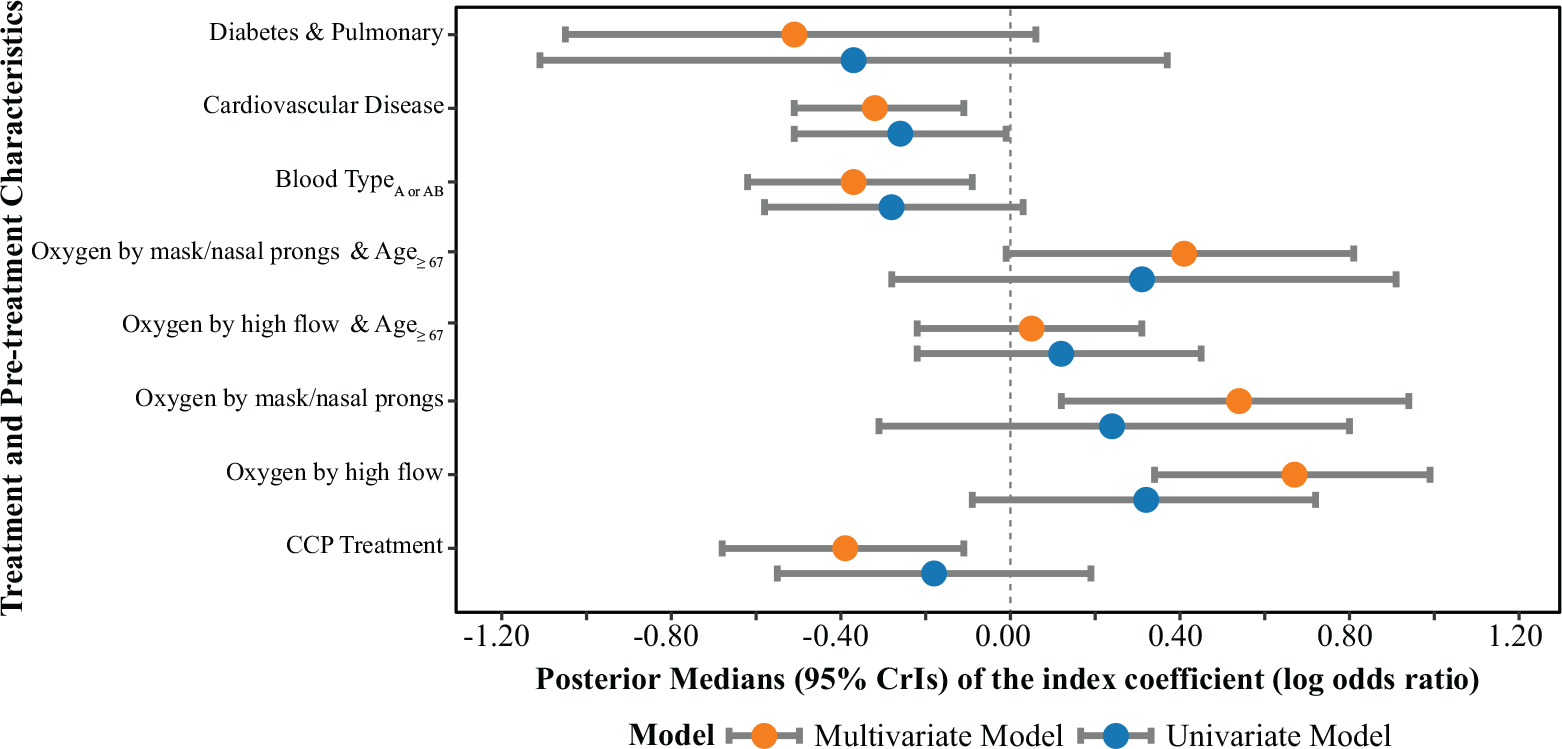
Comparison of univariate and multivariate models with respect to posterior distributions of index coefficients, summarized by the posterior medians and 95% credible intervals, for treatment and pre-treatment characteristics (*log*OR) corresponding to the primary ordinal outcome.

Figure 3 indicates that the multivariate model offers better precision when estimating coefficients for pre-treatment characteristics and treatment in comparison to the univariate model, as indicated by narrower 95% CrIs. In the multivariate model, most of the coefficients’ 95% CrIs do not encompass zero. In contrast, for the univariate model, the 95% CrIs for almost all coefficients include zero. These coefficients correspond to the *log*OR associated with the pre-treatment characteristics for the primary ordinal outcome, and by convention, if a 95% CrI of *log*OR covers zero, we do not draw a definitive conclusion regarding whether patients with these pre-treatment characteristics would benefit more from CCP than from control treatment.

As indicated in Table 1, the findings from the multivariate model are consistent with the results reported by Park et al. [24]: patients with pre-existing conditions, such as cardiovascular (posterior median of OR = exp(−0.32) = 0.73), diabetes, and pulmonary (posterior median of OR = exp(−0.51) = 0.60) diseases, blood type A or AB (posterior median of OR = exp(−0.37) = 0.69), and those at an early stage of COVID-19 (indicated by hospitalized but no oxygen therapy required), are expected to benefit the most from CCP treatment. On the other hand, patients without pre-existing conditions and those at more advanced stages of COVID-19 might potentially experience harm (posterior medians of OR = exp(0.54) = 1.72 and OR = exp(0.67) = 1.95). In addition, the proposed Bayesian model provides lower levels of uncertainty in the estimation of the ORs.

For each patient, the effect of CCP treatment versus control on each outcome, as measured by OR, is calculated based on the patient’s pre-treatment characteristics and the posterior distributions of coefficients derived from either the multivariate or the univariate model. The TBI is subsequently computed in accordance with Equation (4). Figure 4 presents a side-by-side comparison of the fitted models, illustrating the relationship between the TBI and the posterior mean of the OR for different outcome types in COMPILE. The left plot is based on the proposed multivariate model (6), in which the x-axis represents the TBI. The right plot is based on the univariate model (7). An odds ratio for CCP efficacy below 1 (dashed grey horizontal lines) indicates a more favorable outcome with CCP treatment compared to the control treatment, and the degree of treatment benefit from CCP is monotonically parameterized by the TBI.

**Fig. 4.**
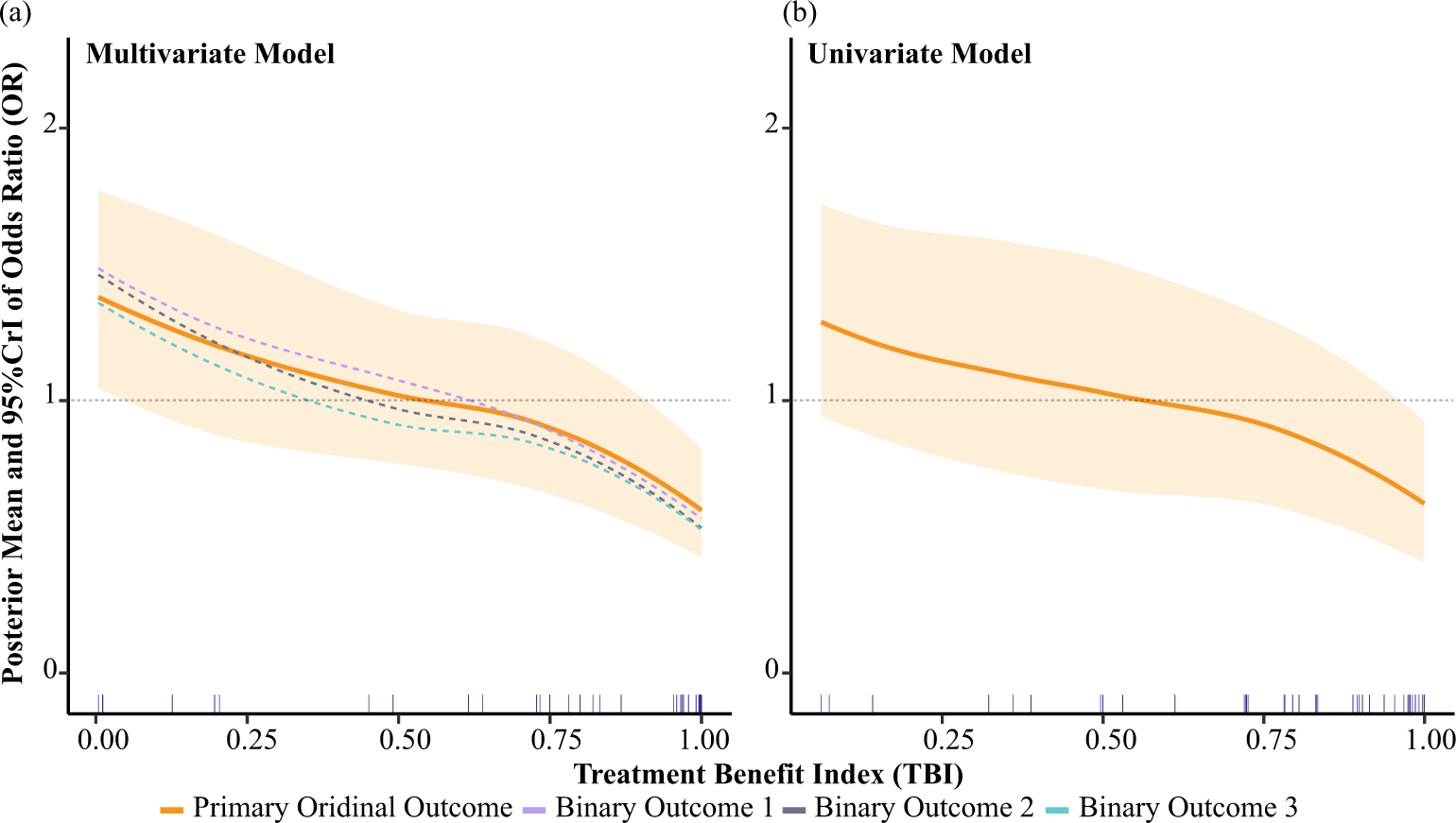
Posterior distributions of odds ratios (ORs) associated with the treatment benefit index (TBI) using the multivariate model (left plot) and univariate model (right plot). In each plot, the solid line represents the posterior mean of OR for the primary ordinal outcome, and the colored band represents the 95% credible interval (CrI) of this OR. The dashed curves in (a) correspond to the posterior means of the ORs for three binary outcomes. The supplementary outcomes are as follows: (1) the binary outcome of hospitalization at day 28, (2) the binary outcome of ventilation or worse at day 28, and (3) the binary outcome of mortality at day 28. The loess smoothing method is applied to illustrate the overall trends. Rug plots at the bottom of each plot represent the data density along the x-axis. An odds ratio for COVID-19 convalescent plasma (CCP) efficacy below 1 (dashed grey horizontal line) indicates a more favorable outcome with CCP treatment compared to the control treatment.

A notable observation from Figure 4 is the narrower 95% credible interval of the OR for the primary ordinal outcome when employing the multivariate model (6), compared to the univariate model (7). This suggests that the multivariate model incorporates and reflects richer available information from the multiple outcomes collected in the trial. Consequently, this improved accuracy may contribute to more informed clinical decision-making based on a more reliable representation of the relationship between CCP efficacy and TBI.

We assessed the goodness-of-fit for both the Bayesian multivariate and univariate models, Models (6) and (7), using posterior predictive checking, a method that evaluates the model’s ability to generate replicated data that closely resembles the observed data [42, 48–50]. The Bayesian p-value was employed to measure the model’s fit, with values near 0.5 suggesting a satisfactory fit. A detailed explanation of the procedure and the results of posterior predictive checking for both the Bayesian multivariate and univariate models is provided in Additional file 5. The results show that both models fit the data well.

## 4 Discussion and conclusions

The current study presents a robust framework for jointly modeling correlated mixed types of health outcomes, which leads to improved precision in estimating heterogeneous treatment effects and optimal ITRs. Our proposed Bayesian multivariate model leverages hierarchical modeling and carefully selected prior distributions to effectively “borrow information” across outcomes, enhancing the estimation accuracy. Through extensive simulations, we compared the proposed model to a Bayesian univariate model, demonstrating that the proposed approach reduces the likelihood of making erroneous optimal ITRs. In the application to an international COVID-19 treatment trial, the proposed model exhibited superior precision in estimating coefficients of treatment and pre-treatment characteristics, as well as in estimating the OR for the primary ordinal outcome. This enables more informed clinical decision-making and highlights the practical applicability of our model in real-world settings.

Our study should be interpreted considering two potential limitations. First, the framework is constrained to situations where the treatment effects and interaction effects across outcomes are positively correlated and maintain a similar scale. When these effects are negatively correlated and possess substantially different scales, our method would need to be adapted to account for such negative associations and disparate scales of effects among outcomes. One potential solution is to use the ideas of group factor analysis [51, 52] to model both positive and negative relationships among outcomes by modeling the residuals as linear transformations of latent factors. Second, the pre-treatment characteristics used for model fitting come from [24], representing the optimal variable set determined through cross-validation. Although no other variable selection method is used in our study, we assessed the model’s goodness-of-fit using posterior predictive checking. The results show that our model fits the data well, suggesting that the direct adoption of pre-treatment characteristics from [24] does not pose a serious limitation. When there is a definite expectation that specific pretreatment characteristics will impact the outcome, those pre-treatment characteristics should be included in the model. When it’s unclear whether certain pre-treatment characteristics should be included, data-dependent variable selection methods [53–57] can be more generally incorporated to potentially improve the current multivariate model.

To the best of our knowledge, no previous studies have jointly modeled mixed types of outcomes to develop ITRs. Our translatable framework has the potential to efficiently leverage information from multiple health outcomes, making it a valuable tool for not only developing ITRs for COVID-19 but also for various other diseases.

## Data Availability

All data produced in the present study are available upon reasonable request to the authors

## Declarations

### Consent for publication

Not applicable.

## Acknowledgements

The authors acknowledge Rebecca Anthopolos for her guidance in writing the manuscript.

## Abbreviations

ITRs: individualized treatment decision rules
TBI: treatment benefit index
COVID-19: Coronavirus disease 2019
COMPILE: COntinuous Monitoring of Pooled International Trials of ConvaLEscent Plasma for COVID-19 Hospitalized Patients
RCT(s): randomized controlled trial(s)
CCP: COVID-19 convalescent plasma
WHO: World Health Organization
HMC: Hamiltonian Monte Carlo
*co*: cumulative proportional odds
OR(s): odds ratio(s)
MVN: multivariate normal distribution
AUC: area under the curve
ROC: receiver operating characteristic
PCD: proportion of correct decisions
IQR: interquartile range
SD: standard deviation
CrI: Credible Interval

## Availability of data and materials

The datasets used and analyzed during the current study are available from the corresponding author upon reasonable request.

## Ethics approval and consent to participate

Not applicable.

## Authors’ contributions

DW: Engaged in problem discussions, contributed to the development of Bayesian hierarchical models, conducted all simulations, and wrote the initial manuscript. KSG, EP, and HGP: Provided supervision, project management, and funding acquisition, participated in problem discussions, contributed to the development of Bayesian hierarchical models, and reviewed and revised the manuscript draft.

All authors read and approved the final manuscript.

