## Supplemental materials for "Improving Individualized Treatment Decisions: A Bayesian Multivariate Hierarchical Model for Developing a Treatment Benefit Index using Mixed Types of Outcomes" (from another version of this preprint: version 1, 2023-11-18)

### Supplementary Materials

Additional file 1 — Stan code for the Bayesian multivariate hierarchical model

```
data {
  int<lower=0> N;                 // number of observations
  int<lower=0> D;                 // number of of binary outcomes
  int<lower=2> L;                 // number of WHO categories
  int<lower=0> P_main;            // number of pre-treatment characteristics in the main effects term
  int<lower=0> P_inter;           // number of pre-treatment characteristics in the interaction effects term
  int<lower=1,upper=L> y_ord[N]; // vector of ordinal outcomes
  int<lower=0,upper=1> y_b[N,D]; // matrix of D binary outcomes (N x D matrix)
  int<lower=0,upper=1> A[N];      // treatment or control
  row_vector[P_main] x_main[N];  // pre-treatment characteristics in the main effects term  (N x P_main matrix)
  row_vector[P_inter] x_inter[N];// pre-treatment characteristics in the interaction effects term  (N x P_inter matrix)
}

parameters {
  ordered[L-1] tau;               // cut-points for cumulative odds model
  vector<lower=0>[(P_inter + 1)] sigma_beta; // sd of outcome-specific treatment main effect and interaction effect
  vector[D] beta_0;               // outcome-specific intercepts for D binary outcomes
  matrix[P_main,(D + 1)] beta_1; // covariates main effect for D binary outcomes and 1 ordinal outcome (P_main x (D + 1) matrix)
  vector[(P_inter + 1)] beta_star; // pooled treatment main effect and pooled interaction effect across all outcomes

  // non-central parameterization

  matrix[(P_inter + 1),(D + 1)] z_beta_int;
}

transformed parameters {
  matrix[(P_inter + 1),(D + 1)] beta_int; // outcome-specific treatment main effect and interaction effect ((P_inter + 1) x (D + 1) matrix)
  vector[D] yhat_b[N];
  real yhat_ord[N];

  for (j in 1:(P_inter + 1))
     for (k in 1:(D + 1)){
        beta_int[j,k] = beta_star[j] + sigma_beta[j] * z_beta_int[j,k];
  }

  for (i in 1:N){
```

```
    for (k in 1:D){
      yhat_b[i,k] =  beta_0[k] + x_main[i] * beta_1[,k]  +  (
    append_col(1, x_inter[i]) * beta_int[,k]) * A[i];
    }
    yhat_ord[i] = x_main[i] * beta_1[,D+1]  +  (append_col(1, x_inter[
    i]) * beta_int[,D+1]) * A[i];
  }
}


model {

  // priors

  sigma_beta ~ exponential(1);
  beta_star ~ normal(0,2.5);
  to_vector(beta_1) ~ normal(0,2.5);
  to_vector(z_beta_int) ~ std_normal();

  for (l in 1:(L-1)){
    tau[l] ~ student_t(3,0,8);
  }

   for (k in 1:D){
    beta_0[k] ~ student_t(3,0,8);
  }

  // outcome model

  for (i in 1:N){
    y_ord[i] ~ ordered_logistic(yhat_ord[I], tau);
    for (k in 1:D){
      y_b[i,k] ~ bernoulli_logit(yhat_b[i,k]);
    }
  }
}
```

Additional file 2 — Main analysis: comparing the performance of the Bayesian multivariate and univariate models when the true ITR is determined by potential outcomes

To implement this potential outcomes-based ITR, we first consider the patient characteristics $\tilde{\boldsymbol{x}}_i$ along with the true values of parameters from the data generation process. Next, we use the *simstudy* package [43] to generate potential primary ordinal outcomes for subjects receiving the control treatment $(y^{(1)}_{A=0})$ and the experimental treatment $(y^{(1)}_{A=1})$. The optimal ITR is derived from the indicator function $I(y^{(1)}_{A=1} < y^{(1)}_{A=0})$, which evaluates whether the experimental treatment outcome is better than the control treatment outcome.

Utilizing this new potential outcomes-based ITR, the subsequent plot (Figure A1) illustrates the comparison of PCD and AUC values between the Bayesian multivariate and univariate models across varying training set sizes. In

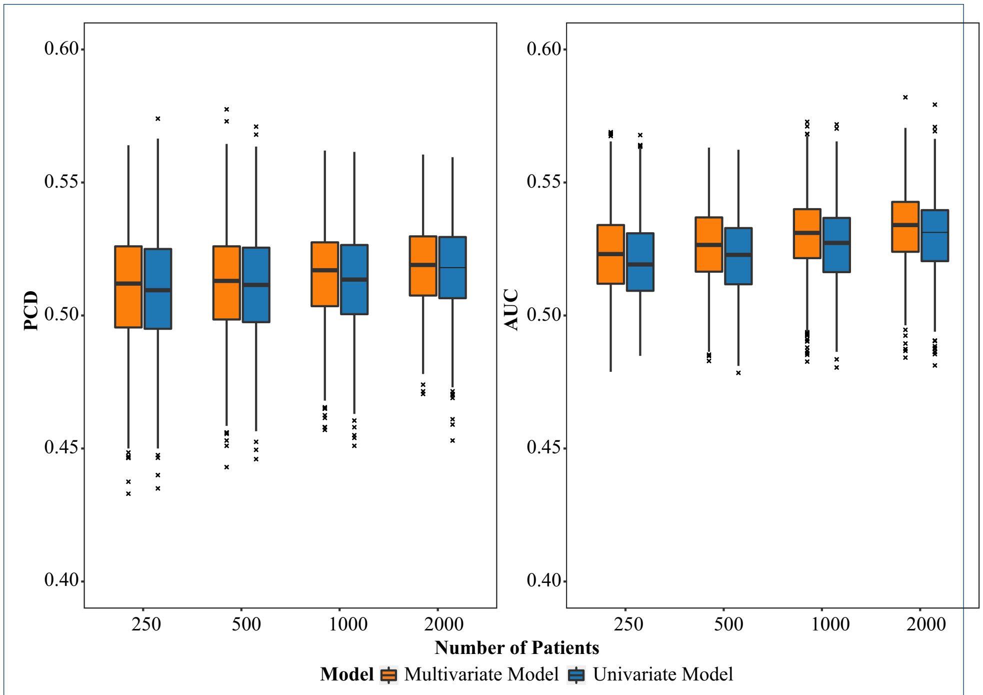


**Figure A1** Utilizing the new potential outcomes-based ITR: the boxplots of PCD and AUC in the test sets, comparing the multivariate (orange) and the univariate (blue) models across different training set sizes (as indicated in the x-axis).

comparison to Figure 1, the improvement in prediction using the multivariate model is less remarkable. This can be attributed to the fact that generating potential outcomes based on probability inherently involves more randomness. The gain in estimation is relatively small compared to the magnitude of this randomness. Consequently, when considering prediction error, the improvement becomes less noticeable as it is overshadowed by the noise introduced by the randomness.

Additional file 3 — Sensitivity analysis: comparing the performance of the Bayesian multivariate and univariate models when the true ITR is determined by potential outcomes

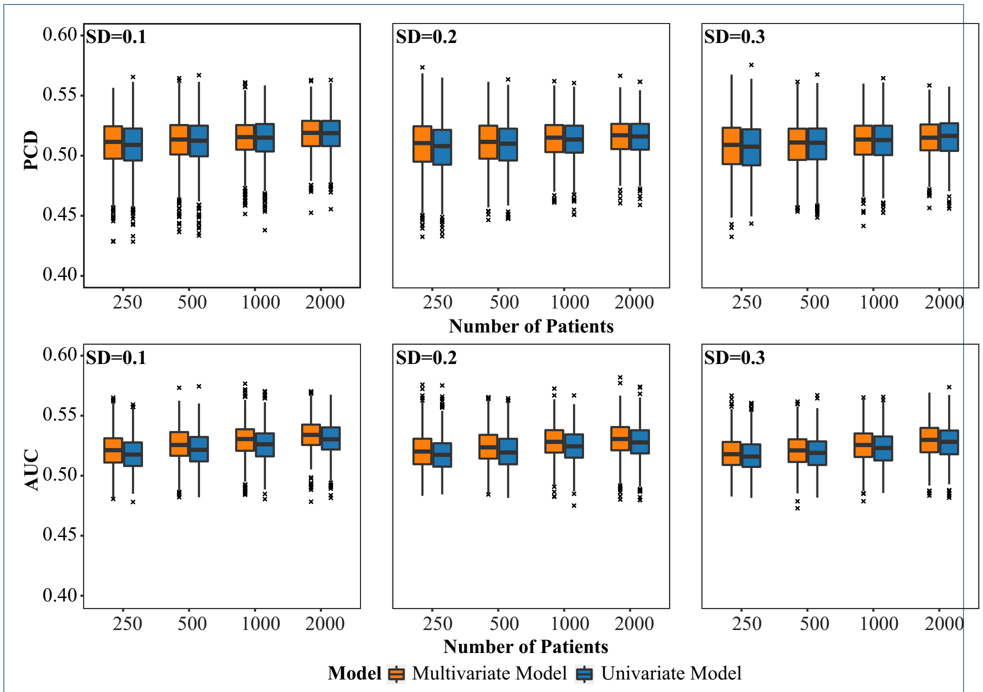


**Figure A2** Utilizing the new potential outcomes-based ITR and different SD for random effects in data generation: the boxplots of PCD and AUC in the test sets, comparing the multivariate (orange) and univariate (blue) models across different training set sizes (as indicated in the x-axis). Three different levels of SD for random effects are considered in data generation: SD=0.1, SD=0.2, and SD=0.3.

Compared to Figure 2, the utilization of this new potential outcomes-based ITR yields less remarkable improvement in the multivariate model's performance. This could be due to the probabilistic nature of generating potential outcomes, which inherently involves more randomness. Despite the gain in estimation, the magnitude of this randomness is relatively large, resulting in a small improvement that is overshadowed by the introduced noise when considering prediction error.

661 Additional file 4 — The WHO 11-point COVID-19 clinical status scale.

| | |
|---|---|
| 0: | Uninfected, no viral RNA detected |
| 1: | Asymptomatic, viral RNA detected |
| 2: | Symptomatic, independent |
| 3: | Symptomatic, assistance needed |
| 4: | Hospitalized, no oxygen therapy |
| 5: | Hospitalized, oxygen by mask or nasal prongs |
| 6: | Hospitalized, oxygen by non-invasive ventilation or high flow |
| 7: | Intubation & mechanical ventilation, $pO_2/FiO_2 \geq 150$ (or $SpO_2/FiO_2 \geq 200$)[a] |
| 8: | Mechanical ventilation, $pO_2/FiO_2 < 150$ (or $SpO_2/FiO_2 < 200$) or vasopressors |
| 9: | Mechanical ventilation, $pO_2/FiO_2 < 150$ and vasopressors, dialysis, or ECMO[b] |
| 10: | Dead |

[a] $pO_2$: partial pressure of oxygen, $FiO_2$: fraction of inspired oxygen, $SpO_2$: oxygen saturation.
[b] ECMO: extracorporeal membrane oxygenation.

**Table A1** The WHO 11-point COVID-19 scale definition[39].

| | **Control (n = 1097)** | **CCP (n = 1190)** |
|---|---|---|
| WHO = 0 | 114 | 150 |
| WHO = 1 | 151 | 168 |
| WHO = 2 | 365 | 386 |
| WHO = 3 | 134 | 142 |
| WHO = 4 | 45 | 45 |
| WHO = 5 | 86 | 84 |
| WHO = 6 | 29 | 52 |
| WHO = 7 | 23 | 30 |
| WHO = 8 | 23 | 36 |
| WHO = 9 | 33 | 22 |
| WHO = 10 | 94 | 75 |

**Table A2** The number of patients at different clinical stages of COVID-19 measured on the WHO 11-point scale at day 14 by treatment group.

Additional file 5 — Goodness-of-fit using posterior predictive checking

In evaluating the suitability of a statistical model, it is crucial to determine whether the model provides an accurate representation of the observed data. This is particularly important for models like the $co$ model, which is built upon a strong assumption of proportional cumulative odds. Posterior predictive checking serves as an effective way of assessing a model's *goodness-of-fit* [47, 48]. This method operates on the premise that a well-fitted model should enable the generation of replicated data ($D^{\text{rep}}$) that resembles the observed data ($D^{\text{original}}$) [49].

The lack of fit can be measured by the Bayesian p-value, which represents the probability of the test statistic (e.g., $P(Y \leq y), y = 0, \ldots, 9$) for $D^{\text{rep}}$ being equal to or exceeding the test statistic for $D^{\text{original}}$. A Bayesian p-value approaching zero or one signifies a potential issue with the model's fit, whereas a value near 0.5 suggests that the model captures the data well [42, 57]. We employed the procedure outlined in [41] to examine the $co$ model's fit to the observed data and to compute the Bayesian p-value.

Table A3 provides the results of posterior predictive checking based on ten test statistics (along with their 95% CrIs) for both the multivariate model (6) and univariate model (7). Our analysis confirmed the satisfactory fit of both models to the data.

| Treatment | **Control** | | | **CCP** | | |
|---|---|---|---|---|---|---|
| Test quantity: % subjects | $T(D^{\text{original}})$ | 95% int. for $T(D^{\text{rep}})$ | Bayesian P value | $T(D^{\text{original}})$ | 95% int. for $T(D^{\text{rep}})$ | Bayesian P value |
| **Multivariate model** | | | | | | |
| WHO $\leq 0$ | 10.39 | [8.39, 12.94] | 0.55 | 12.61 | [10.08, 14.71] | 0.39 |
| WHO $\leq 1$ | 24.16 | [20.51, 26.80] | 0.34 | 26.72 | [23.28, 29.58] | 0.41 |
| WHO $\leq 2$ | 57.43 | [52.42, 59.43] | 0.19 | 59.16 | [55.71, 62.52] | 0.48 |
| WHO $\leq 3$ | 69.64 | [65.36, 71.93] | 0.29 | 71.09 | [68.24, 74.45] | 0.55 |
| WHO $\leq 4$ | 73.75 | [69.92, 76.12] | 0.32 | 74.87 | [72.44, 78.32] | 0.63 |
| WHO $\leq 5$ | 81.59 | [78.21, 83.68] | 0.34 | 81.93 | [80.17, 85.29] | 0.74 |
| WHO $\leq 6$ | 84.23 | [82.22, 87.15] | 0.64 | 86.30 | [83.78, 88.40] | 0.44 |
| WHO $\leq 7$ | 86.33 | [84.78, 89.43] | 0.74 | 88.82 | [86.13, 90.50] | 0.33 |
| WHO $\leq 8$ | 88.42 | [87.69, 91.89] | 0.90 | 91.85 | [88.82, 92.69] | 0.14 |
| WHO $\leq 9$ | 91.43 | [90.43, 94.07] | 0.81 | 93.70 | [91.26, 94.71] | 0.22 |
| **Univariate model** | | | | | | |
| WHO $\leq 0$ | 10.39 | [8.57, 13.13] | 0.62 | 12.61 | [9.83, 14.54] | 0.32 |
| WHO $\leq 1$ | 24.16 | [20.78, 27.07] | 0.41 | 26.72 | [23.03, 29.41] | 0.36 |
| WHO $\leq 2$ | 57.43 | [52.42, 59.80] | 0.23 | 59.16 | [55.55, 62.44] | 0.46 |
| WHO $\leq 3$ | 69.64 | [65.45, 72.20] | 0.31 | 71.09 | [68.15, 74.45] | 0.55 |
| WHO $\leq 4$ | 73.75 | [69.83, 76.30] | 0.34 | 74.87 | [72.44, 78.40] | 0.62 |
| WHO $\leq 5$ | 81.59 | [78.12, 83.87] | 0.35 | 81.93 | [80.17, 85.29] | 0.74 |
| WHO $\leq 6$ | 84.23 | [82.04, 87.24] | 0.64 | 86.30 | [83.78, 88.49] | 0.45 |
| WHO $\leq 7$ | 86.33 | [84.69, 89.52] | 0.74 | 88.82 | [86.13, 90.50] | 0.34 |
| WHO $\leq 8$ | 88.42 | [87.60, 91.89] | 0.89 | 91.85 | [88.82, 92.69] | 0.14 |
| WHO $\leq 9$ | 91.43 | [90.34, 94.17] | 0.81 | 93.70 | [91.34, 94.79] | 0.23 |

**Table A3** Summary of posterior predictive checking based on the ten test statistics.
